# Problems bring along their solutions: Characterization of a carbapenem-resistant *Citrobacter braakii* strain and its lytic phage both isolated from sewage water

**DOI:** 10.1101/2025.05.09.25327349

**Authors:** Johanna Pukall, Wencke Rode, Johanna Albrecht, Simon Oerters, Rabea Schlüter, Tjorven Hinzke, Daniela Zühlke, Susanne Sievers

**Author notes:** correspondence to Susanne Sievers, and Daniela Zühlke.

## Abstract

*Citrobacter* spp. are facultative anaerobic, Gram-negative bacteria that have been known to cause infections in the urinary tract and bloodstream. These infections are predominantly nosocomial in nature. Hospitals are considered to be reservoirs for multiresistant bacteria; as such, their wastewater microbiome is characterized by these bacteria. In the present study, a carbapenem-resistant *Citrobacter braakii* strain was isolated from hospital sewage in Greifswald, Germany. This particular strain, designated *C. braakii* GW-Imi-1b1, was the focus of a comprehensive investigation including analyses of antibiotic resistances and proteome profile. Antibiotic resistance tests revealed a broad resistance spectrum, including chloramphenicol, tetracycline, and ertapenem. In addition, an investigation into the defense mechanisms revealed a constitutive presence of a β-lactamase at an unusually high concentration. The growth curve with different meropenem concentrations revealed almost no effect on growth at 20 µg/ml meropenem. Consequently, the high prevalence of β-lactamase may be a contributing factor to the observed carbapenem resistance. Furthermore, due to its localization on the plasmid, it may be capable of being transferred within the wastewater microbiome. Antibiotic treatment of bacterial infections can be substituted or accompanied by phage therapy. The lytic phage vB_CbrP_HGW_001 was isolated from sewage and has been shown to be capable of lysing *C. braakii* GW-Imi-1b1. Sequencing and microscopic analysis revealed a *Kayfunavirus* within the *Caurovicetes.* Subsequent properties of vB_CbrP_HGW_001 indicated the potential for a promising alternative treatment to antibiotics. Phage therapy, particularly when employed in combination with antibiotics, demonstrates a valuable tool in the combat against infections caused by multidrug-resistant bacteria.

## 1 Introduction

*Citrobacter* spp. are facultative anaerobic, Gram-negative bacteria from the order *Enterobacterales*. They are commonly found in various environments, including water, soil, food, and the intestinal tracts of animals and humans and they are generally not considered to be prevalent pathogens (1,2). However, it has been documented that some species cause primarily urinary tract and bloodstream infections, predominantly nosocomial in nature (3). Less commonly, infections of the skin, intra-abdominal region, and respiratory tract have been reported (4).

*C. freundii* is the most common pathogen, but other *Citrobacter* species such as *C. koseri* and *C. braakii* can also cause infections (4). Especially individuals with a compromised immune system and neonates are suspectible by pathogenic *Citrobacter* spp. (5). For instance, a bloodstream infection caused by *C. braakii* was reported in an immunocompromised allogeneic stem cell transplant patient (6). An analysis of nine newborns diagnosed with *C. freundii* sepsis, of which one infant died after 7 h of infection and three developed meningitis, revealed multi-drug resistances of each isolate (7). Antimicrobial resistances (AMR), especially multi-drug resistance, represent a major global health care problem in *Citrobacter* spp. and beyond (3).

A systematic review and meta-analysis examined the epidemiology of *Citrobacter* spp., revealing increased multidrug-resistance in species causing infections in hospitalized patients (3). Hence, it is of utmost importance to further characterize *Citrobacter* pathogens and their resistance mechanisms and to come up with novel treatment strategies to fight AMR infections. The opportunistic pathogen *C. braakii* GW-Imi-1b1, analyzed in this study, was isolated from hospital sewage water in Greifswald (Germany). Hospitals provide an ecological niche and select for for antibiotic-resistant bacteria, i.e. the wastewater of hospitals contains bacteria that harbor various resistances (8).

*C. braakii* GW*-*Imi-1b1 was detected in the inhibition zone of an imipinem-sensitive *C. portucalensis* isolate (9). Imipinem belongs to the carbapenems, a class of β-lactam antibiotics with one of the broadest antibacterial activities for use in humans (10). They are resistant to hydrolysis by the majority of β-lactamases and are typically employed in the treatment of severe nosocomial infections caused by multi-resistant bacteria, such as ESBL-producing *Enterobacteriaceae* and *Pseudomonas aeruginosa* (11). Therefore it is not surprising that, according to the WHO, *Enterobacterales* resistant against this last-resort antibiotic belong to the most threatening bacteria to human health (12). This resistance refers to several resistance mechanisms. Mostly carbapenem-resistant *Enterobacterales* harbor carbapenamases, which are β-lactamases capable of hydrolyzing carbapenems. Additionally the penicillin-binding protein can be modified, thereby impeding its interaction with carbapenems and thus its antibacterial effects. Furthermore, altered pore proteins and an increased expression of the efflux pump, typically the AcrAB-TolC efflux pump, lead to a decreased import and increased export of carbapenems. Lastly, an alteration of biofilm structure has been demonstrated to induce drug resistance (13).

Since there are only limited treatment options against multiresistant bacteria available, alternative therapies like phage therapy should be considered more broadly. Bacteriophages are viruses infecting bacteria and were already discovered in 1917 by Félix Hubert d’Herelle (14). They are characterized by a considerable degree of genomic and morphological diversity and can be classified into two categories: tailed and non-tailed virions. Furthermore, the genomes of phages can be either double-stranded or single-stranded DNA or RNA (15). However, the most prevalent phages generally contain double-stranded DNA and belong to the tailed phages. Of particular significance within the context of phage therapy against multiresistant bacteria are lytic phages. Most phages feature a narrow host range bringing the advantage that they specifically infect only their target host bacterium but leave other bacteria within a microbiome unaffected. Since lytic phages kill their host at the end of their replication, they are suitable for therapy (16). Another replication strategy alongside the lytic is the lysogenic replication. Instead of the lysis of the host following the assembly of newly synthesized bacteriophages, the bacteriophage genome is integrated into the bacterial genome or maintained as an episomal element. Subsequently, the viral genomic information undergoes replication within the host’s genome. However, lysogenic phages might become lytic in response to environmental changes (17).

Despite the fact that phages had been discovered over a century prior, the introduction of antibiotics led to a stagnation of phage therapy research in the Western world. Given the emergence of multiresistant bacteria as a global health concern, phages have resurged as a promising alternative treatment option (18).

This study characterizes the aforementioned carbapenem-resistant *C. braakii* GW-Imi-1b1, which was already sequenced by Schneider *et al.* in 2023 (9). Mass spectrometric methods and phenotypic analyses were employed to elucidate the proteome under various conditions and strain properties, including the antibiotic profile, respectively. Notably, a lytic bacteriophage against *C. braakii* GW-Imi-1b1 was also isolated from sewage water and further characterized, and could serve as an alternative treatment option against this carbapenem resistant strain in the clinic.

## 2 Material and Methods

### Strain isolation

*C. braakii* GW-Imi-1b1 was isolated from the hospital sewage of Greifswald in October, 2020 on Klebsiella *ChromoSelect* Selective Agar (Sigma-Aldrich). Antibiotic resistance profiles with a total of 11 antibiotics were evaluated, following the EUCAST recommendations for *Enterobacterales* (19). Based on the carbapanem resistance, *C. braakii* GW-Imi-1b1 was used for further in-depth analysis.

### Genome sequencing

For sequencing, the strain GW-Imi-1b1 was cultured overnight (37 °C, 180 rpm) in 10 ml Mueller Hinton broth (MHB) in glass reaction tubes. Cultures were centrifuged (10,000 xg, 4 °C), the supernatant discarded and cell pellets washed with sterile PBS. Cells were pelleted again under same conditions, the supernatant discarded and pellets stored at −70 °C and shipped on dry ice to the institute of microbiology and genetics in Göttingen, Germany. The genome sequencing results have been published in Schneider *et al.* in 2023; the strain GW-Imi-1b1 was identified as *Citrobacter braakii*.

### Phenotypical characterization of *C. braakii* GW-Imi-1b1

#### Scanning electron microscopy

To prepare the cells for scanning electron microscopy, 1 ml of a mid-exponential phase of a *C. braakii* GW-Imi-1b1 culture was mixed with 9 ml of 0.9 % NaCl and the suspension was filtered through a polycarbonate filter (pore size 0.2 µm; Merck Millipore, Darmstadt, Germany). A piece of the filter was transferred to 1 ml of the fixative solution (20 mM HEPES, 1 % glutaraldehyde, 4 % paraformaldehyde, 0.2 % picric acid), incubated for 1 h at room temperature (RT) and then stored overnight at 4 °C. Samples were washed with washing buffer (100 mM cacodylate buffer [pH 7], 1 mM CaCl_2_) three times for 10 min each time, treated with 2 % tannic acid in washing buffer for 1 h at RT, and washed again with washing buffer three times for 15 min each time. After that, samples were dehydrated in a graded series of aqueous ethanol solutions (10 %, 30 %, 50 %, 70 %, 90 %, 100 %) on ice for 15 min each time. Before the final change of 100 % ethanol, samples were allowed to reach RT and then critical point-dried with liquid CO_2_. Finally, samples were mounted on aluminum stubs, sputtered with gold/palladium and examined with a field emission scanning electron microscope Supra 40 VP (Carl Zeiss Microscopy Deutschland GmbH, Oberkochen, Germany) using the Everhart-Thornley SE detector and the in-lens detector in a 80:20 ratio at an acceleration voltage of 5 kV. All micrographs were edited by using Adobe Photoshop CS6.

#### Resistance profile

Antimicrobial susceptibility testing of *C. braakii* GW-Imi-1b1 was performed using the disc diffusion method, also known as the Kirby-Bauer method, according the EUCAST guidelines (19). Briefly, the strain was cultured in MHB overnight and cell density was adjusted to a 0.5 McFarland turbidity standard. The cell suspension was inoculated on MH agar plates using sterile cotton swabs. Antimicrobial discs (bestbion dx GmbH, Hürth, Germany) were placed firmly on the surface of the inoculated agar plate. Agar plates were incubated upside-down at 37°C for 24 h. Subsequently, inhibition zones were measured and interpreted according to the EUCAST breakpoints. The following antibiotic discs were used: amikacin (AK) 30 µg, amoxicillin + clavulanic acid (AUG) 30 µg, aztreonam (ATM) 30 µg, cefotaxime (CTX) 5 µg, ceftazidime (CAZ) 10 µg, chloramphenicol (C) 30 µg, ciprofloxacin (CIP) 5 µg, ertapenem (ETP) 10 µg, imipenem (IMI) 10 µg, levofloxacin (LEV) 5 µg, meropenem (MRP) 10 µg, ofloxacin (OFX) 5 µg, tetracyclin (TE) 30 µg, tobramycin (TOB) 10 µg, trimethoprim/sulfamethoxazole (SXT) 25 µg, cefepime (CEP) 30 µg.

Epsilometertests (E-test) for determination of minimal inhibitory concentration (MIC) of carbapenem antibiotics were done in the same fashion as disc assays on MH agar plates using MIC test strips for ertapenem, imipenem, and meropenem, with a concentration ranging from 0.002-32 µg/ml (bestbion dx GmbH, Hürth, Germany).

#### Characterization of pathogenicity factors

To screen for potential pathogenicity factors of *C. braakii* GW-Imi-1b1, motility assays and a protease assay were conducted. All assays were also performed with the control strains *Pseudomonas aeruginosa* PAO1 (20).

Swimming and swarming motility was tested. For each assay, overnight cultures were prepared by inoculating 5 ml LB and incubating overnight at 37 °C at 150 rpm. To assess the swimming motility, 5 µl of cell culture was positioned at the center of the specific agar (10 g/l trypton, 3 % agar, 5 g/l NaCl) and subsequently incubated at 37 °C for 12-14 h. Swarming motility was tested just as swimming motility, but using a different semi-solid medium (8 g/l LB, 0.5 % glucose, 0.5 % agar) and subsequent incubation at 30 °C for 12-14 h. Motility was documented in pictures after the respective incubation time.

Protease secretion was examined in different growth phases. For that, LB medium was inoculated to a OD_540nm_ of 0.08 from a previously prepared 5 ml LB overnight culture, and the bacteria incubated at 37 °C and 150 rpm. Cells were harvested at the exponential (t1: OD_540nm_ 0.5), transient (t2: OD_540nm_ 1), early stationary (t3: OD_540nm_ 2), and late stationary phase (t4: OD_540nm_:3-4). The culture was sterile filtered through a 0.2 µm filter and subsequently used for protease activity assays. In addition to the previously mentioned positive control, the corresponding quorum-sensing-deficient mutant *Pseudomonas aeruginosa* Δ*lasI/rhII* (21) was used as a negative control. Protease activity assays were based on azocasein cleavage detection. To this end, an azocasein solution was prepared composed of 50 mM Tris, 5 mM EDTA and 1 % azocasein and adjusted to a pH of 7.5. Some 150 µl of the respective sterile filtered culture medium were mixed with 250 µl of the azocasein solution and incubated at 37 °C for 6 h. Some 1.2 ml of 10 % TCA were added and again incubated for 25 min at room temperature. Following centrifugation for 15 min at 21,382 xg, 600 µl of the sample were mixed with 750 µl of 1 M NaOH in a cuvette. Absorption was subsequently measured at 440 nm. For reference, a negative sample consisting of 250 µl of azocasein solution, 150 µl of LB and 1.2 ml of TCA were mixed and 600 µl of the mixture was added to 750 µl of 1 M NaOH. The protease activity is proportional to the intensity of the dye measured at an extinction of 440 nm and divided by OD_500nm_ of the respective time point of harvest.

### Proteome analysis of *C. braakii* GW-Imi-1b1

To characterize the protein profile of *C. braakii* GW-Imi-1b1, extra- and whole cell proteins were extracted and analyzed. For that, the strain was cultivated in LB to stationary phase (OD_540nm_ 1.5). Subsequently, 30 ml of the culture were harvested and centrifuged for 10 min at 4 °C and 9,693 xg. For cellular protein extraction, the resulting pellet was washed twice in 1 ml TE-Buffer (10 mM Tris, 1 mM EDTA, pH 7.5) and subsequently resuspended in 1 ml TE-Buffer with 1% Triton-X. The suspension was transferred into a 2 ml screw cap micro tube with 500 mg 0.1 mm beads and mechanically disrupted three times with a FastPrep-24TM5G (MP Biomedicals, Irvine, USA) at 6.5 m/s for 30 sec. After centrifugation for 5 min at 4 °C and 21,382 xg, the supernatant was transferred into a new reaction tube and again centrifuged. Subsequently, −20°C cold acetone equal to six times the volume of the supernatant was added, and the resulting solution was incubated overnight at −20 °C. On the next day, the sample was centrifuged again for 1 h at 4 °C and 21,382 xg. The precipitated proteins were washed in 1 ml aceton and subsequently dried in a vacuum centrifuge. The dried pellet was resuspended in 150 µl of 8 M urea and 2 M thiourea under shaking at 150 rpm. Samples were stored at −20 °C until further processing.

Extracellular proteins were precipitated from the supernatant after harvesting with TCA. Briefly, 3.3 ml 100 % TCA were added to 30 ml supernatant and incubated overnight at 4 °C. Samples were then centrifuged for 1 h at 4 °C and 9,693 xg. The resulting protein pellet was dissolved in 1 ml of 70 % ethanol and again centrifuged for 5 min at room temperature and 16,060 xg in a new reaction tube. This washing step was repeated once with 70 % ethanol and 100 % ethanol. Subsequently, protein pellets were dried in a vacuum centrifuge and resuspended in 50 ml 8 M urea and 2 M thiourea. Samples were stored at −20 °C until further processing. Protein concentration was determined with ROTI^®^ Nanoquant (Carl Roth GmbH, Karlsruhe, Germany) according to the manufacturers protocol.

### Effect of meropenem on the Citrobacter spp. protein profile

For pre-culturing, 100 ml MHB in 500 ml Erlenmeyer flasks were inoculated with one colony from 24 h incubated (37 °C) MH agar plates of *C. braakii* GW-Imi-1b1 and incubated overnight (37 °C, 180 rpm). Material of these cultures was used to inoculate fresh MHB to a final OD of 0.1, and meropenem stock solution (5 mg/ml in PBS) was added to all but the control flasks to reach final concentrations of 0.064, 0.125 or 20 µg/ml meropenem (final volume 100 ml in 500 ml Erlenmeyer flasks). Cultures were incubated at 37 °C and 180 rpm, and samples for proteomics taken from the pre-cultures at the start of the experiment (t0), as well as from flasks after 2 h (t1) and 24 h (t2) incubation. Samples (1.5 ml each) were washed once with PBS (10.000 xg, 4 °C) and the pellets stored at −70 °C until further processing. Additionally, bacterial growth was measured by determining the optical density OD_500nm_ of the culture. Experiments were performed in biological triplicate.

Proteins were extracted from *Citrobacter braakii* GW-Imi-1b1 as previously described (22) with minor modifications. In brief, lysis buffer (50 mM TEAB, 4 % SDS, 50 mM DTT) was added in a 1:5 ratio to pellets, the pellets were carefully resuspended, samples heated (10 min 95 °C) and subsequently sonicated in an ultrasonic bath for 10 min. Samples were cooled on ice in between. Protein concentration was determined with ROTI^®^ Nanoquant (Carl Roth GmbH, Karlsruhe, Germany) according to the manufacturers protocol.

### Protein digestion, mass spectrometry analysis and identification

Digestion of proteins using S-trap micro columns was done as described by Brauer *et al*.

(23). Briefly, 50 µg of protein sample were adjusted to 5 % SDS, proteins reduced with DTT, alkylated with IAA and samples acidified with 12 % phosphoric acid. Proteins were then bound to the S-Trap column and digested with pre-activated trypsin for 3 h at 47 °C. Peptides were eluted with 50 mM TEAB, followed by 0.1 % acetic acid and 60 % acetonitrile with 0.1 % acetic acid. Elutions of the same sample were pooled, dried in a vacuum centrifuge and stored at −70 °C until further processing. The samples were fractionated with Pierce C18 spin columns (ThermoFisher Scientific, Waltham, USA) and application of 18 mg C18 Reprosil Gold (Dr. Maisch HPLC GmbH, Ammerbruch-Entringen, Germany). Peptides were eluted in 8 fractions using increasing amounts of ACN (5 to 50 % ACN) in 0.1 % TEA under the same conditions. Subsequently, we combined fractions 1 and 5, 2 and 6, and so on to generate 4 final fractions. Fractionated samples were transferred to glass vials, dried in a vacuum centrifuge and stored at −80 °C until analysis.

For MS analysis, fractionated samples were resuspended in 10 µl 0.1 % acetic acid and analyzed using an EASY-nLC1200 coupled to a QExactive HF mass spectrometer (Thermo Fisher Scientific, Waltham, USA). Peptides were separated on a self-packed analytical column (100 µm inner diameter, reverse phase C18 material (Dr. Maisch HPLC GmbH, Ammerbruch-Entringen, Germany; 3,6 µm particle size, 20 cm column length) using a 80 min binary non-linear gradient (4% to 50% of 0.1% (v/v) acetic acid in acetonitrile) at a flow rate of 300 nl/min. Survey scan resolution was 60,0000 in a range of 333-1650 m/z and the 15 most-intense peaks per scan were selected for fragmentation. Ions with single charge and ions with unknown charge state were excluded from fragmentation; and precursor ions were dynamically excluded for 30 s from fragmentation. Lock mass correction was enabled (lock mass 445.12003 Da).

Protein identification and label-free quantification was done using MaxQuant software (v1.6.17.0) using a protein sequence database of C. braakii GW-Imi-1b1 (5,338 entries); reverse sequences and common laboratory contaminants were added by the software (24). Protein sequences were generated from the assembled genome and were clustered at 99 % identity using CD-Hit (25) and amended with common laboratory contaminants (ftp://ftp.thegpm.org/fasta/cRAP, version 04.03.2019). The following parameters were set: maximum of 2 missed cleavages, methionine oxidation was set as variable modification, carbamidomethylation of cysteine was set as fixed modification, number of minimal required unique peptides was set to one, match between runs was enabled. Unique and razor peptides were considered for label-free quantification with a minimum ratio count of 2. MaxQuant companion software Perseus was used for analyses of the proteins (26).

The mass spectrometry proteomics data have been deposited to the ProteomeXchange Consortium via the PRIDE partner repository with the dataset identifier PXD063554 (27).

### Lytic phage vB_CbrP_HGW_001

#### Phage isolation of hospital sewage

The lytic phage was isolated from hospital sewage and is designated vB_CbrP_HGW_001. Three ml of the wasterwater was mixed with 30 µl chloroform to eliminate the present bacteria. Some 200 µl of the treated water was then incubated with 200 µl of an overnight culture of *C. braakii* GW-Imi-1b1 for 10 min at room temperature. Present phages were tested by the double agar layer method according to Kropinski *et al* (28). Briefly, 200 µl of the mixture was added to soft agar pre-tempered at 45 °C containing 0.75 % agar in LB medium. After short mixing the content was poured onto base agar containing 1.5 % agar and solidified at room temperature. The plate was subsequently incubated at 37 °C over night.

On the next day, visible plaques were isolated by pricking the top agar layer with a pipette tip. Next, the sample was dissolvedd in 500 µl SM-buffer (0.1 M NaCl, 2.3 mM MgSO_4_) and vortexed for 30 sec. After centrifugation at 14,000 xg at 4 °C for 2 min, the solution was sterile filtered through a 0.2 µm cellulose acetate filter (VWR International). The final lysate was again tested with the already described double agar layer method. This procedure was repeated three times to ensure phage purity. For sequencing, DNA was extracted using Monarch^®^ Mag Viral DNA/RNA Extraction Kit (New England Biolabs). Two ml of a high titer lysate were ultrafiltered using Amicon Ultra 0.5 ml centrifugal filters MWCO 3 kDa (Sigma-Aldrich) for enrichment of the DNA. The resulting 200 µl were used for DNA extraction following the manufacturers protocol. DNA concentration was fluorometrically quantified by Qubit 3.0 Fluorometer (Life Technologies, Carlsbad, California, US) and prepared to a final concentration of 50 ng/µl in 30 µl. Subsequent DNA sequencing was conducted by Eurofins Genomics via Oxford Nanopore Sequencing as part of the Whole Plasmid Sequencing Service.

#### Transmission electron microscopy

Prior microscopy, phage suspension was washed. In brief, 1 ml of a high titer phage suspension was centrifuged at 25,000 xg for 60 min. After washing of the resulting pellet with 1 ml 0.1 M ammoniumacetate (pH 7), phages were dissolved in 300 µl Preissner-buffer (4 mM KH_2_PO_4_, 6 mM K_2_HPO_4_). The flotation method was used for the negative staining procedure. Phages were allowed to adsorb onto a glow discharged carbon-coated holey Pioloform film on a 400-mesh grid for 5 min. The grid was then transferred onto two droplets of deionized water, and finally onto a drop of 1% aqueous uranyl acetate for 15 s. After blotting with filter paper and air-drying, the samples were examined with a transmission electron microscope LEO 906 (Carl Zeiss Microscopy Deutschland GmbH, Oberkochen, Germany) at an acceleration voltage of 80 kV. For acquisition of the images, a wide-angle dualspeed CCD camera Sharpeye (Tröndle, Moorenweis, Germany) was used, operated by the ImageSP software. Finally, all micrographs were edited by using Adobe Photoshop CS6.

#### Phage adsorption assay

The initial characterization of the novel isolated phage was conducted through the examination of its adsorption to its host. The adsorption assay was carried out as described by Wittmann *et al.* (2014) with minor modifications (29). Briefly, exponentially growing cultures of *C: braakii* GW-Imi-1b1 were infected with the phage with a multiplicity of infection (MOI) of 0.01. The infected culture was then further incubated at 37 °C and samples were taken at 5-minute intervals for a duration of 25 min, i. e. 1 ml of the sample was centrifuged at 4,656 xg for 4 min. The titer of free phages was subsequently determined by the following procedure. First, the supernatant was diluted and mixed with an overnight culture of the host in LB soft agar pre-tempered at 45 °C, as previously described. The soft agar was poured on LB-agar and solidified at room temperature. After incubation overnight at 37 °C, resulting plaques were enumerated and the plaque forming units (PFU) were calculated. The adsorption assay was conducted in three biological replicates.

#### Burst size

Burst size experiments were performed as previously described by Morozova *et al* (30) with slight modifications. Some 10 ml of a mid-exponential phase of *C. braakii* GW-Imi-1b1 was centrifuged for 3 min with 4,656 xg and the pellet was resuspended in 500 µl LB medium. The culture was infected with vB_CbrP_HGW_001 at a MOI of 0.01 and incubated for 10 min at 37 °C to ensure attachment to the cell. Then the cells were pelleted again as before, resuspended in 10 ml LB medium and incubated for 40 min. Aliquots of the culture were harvested every 5 min and centrifuged for 4 min at 4,656 xg at 4 °C. The supernatant was used for phage titer determination using the double layer agar method.

#### pH and temperature stability

pH and temperature stability were determined according to Akhwale *et al.* with minor modifications (31). For temperature stability, a phage suspension of 2.5×10^9^ PFU/ml was subjected to various temperatures (21 °C, 30 °C, 37 °C, 40 °C, 50 °C and 60 °C) for 6 h. Thereafter, the samples were cooled on ice, and the titer was determined by the double agar layer method. For testing of the pH stability, 100 µl of the phage suspension was incubated in 900 µl LB with different pH values (2, 4, 6, 8, 10) and subsequently incubated for 3 h at room temperature. Phage titer was again determined by the double agar layer method directly after incubation time.

## 3 Results

### Phenotypical analyses of *C. braakii* GW-Imi-1b1

*C. braakii* GW-Imi-1b1 was visualized by scanning electronmicroscopy, as shown in Figure 1. These rodshaped bacteria measure approximately 2 µm in length and 0.5 µm in width. Interestingly, they apparently produce outer membrane vesicles (OMVs). *C. braakii* GW-Imi-1b1 was further characterized phenotypically including its antibiotic profile, protease secretion ability and motility.

**Figure 1:**
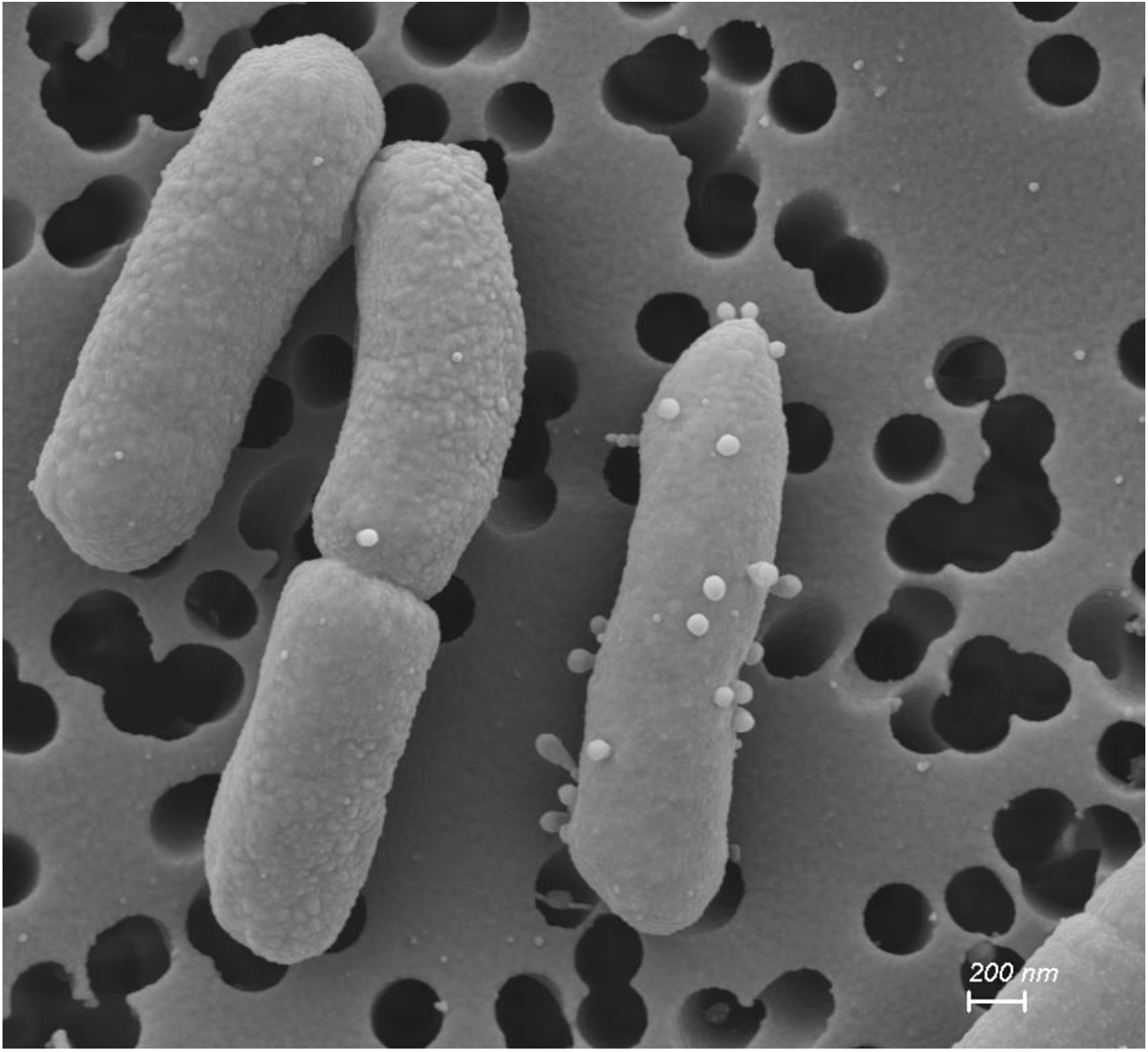
SEM of *C. braakii* GW-Imi-1b1. Cells were harvested in the mid-exponential phase and subsequent applied on 0.2 µm polycarbonate filter.

### Protease secretion

Protease activity was proven by the cleavage of azocasein. In the presence of proteases, the azo dye is cleaved from casein. Consequently, the released dye is quantifiable through photometric analysis. Cleavage can be done by several proteases like serinproteases, metalloproteases and acidproteases (32). As a positive control, *P. aeruginosa* PAO1 known to secrete proteases was used. Notably, protease secretion was observed to occur exclusively at higher cell densities, as no activity was detected in the initial time points. The negative control strain of *P. aeruginosa* Δ*lasI/rhII*, a quorum sensing mutant devoid of protease secretion, exhibited comparable activity to that of *C. braakii* GW-Imi-1b1 indicating that both strains do not secrete proteases.

#### Motility of *C. braakii* GW-Imi-1b1

Swimming and swarming motility was tested and compared with the positive control *P. aeruginosa* PAO1. Images were taken 14 h after application and incubation at 37 °C and are depicted in Figure 3. *C. braakii* GW-Imi-1b1 is incapable of either swarming or swimming motility. A comparison of the size and contour reveals differences to the positive control. The colony morphology of *P. aeruginosa* PAO1 is characterized by a diffuse contour, in contrast to the clearer and smaller forms observed in *C. braakii* GW-Imi-1b1.

**Figure 2:**
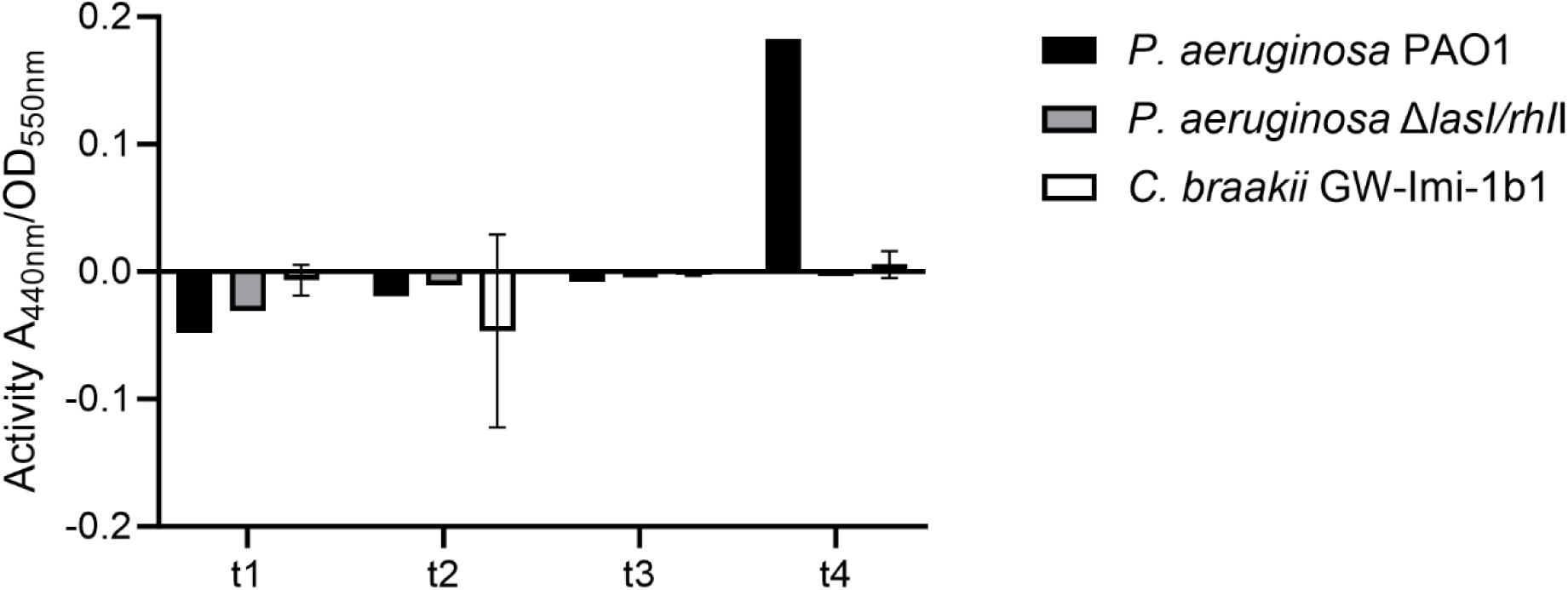
Protease activity of *C. braakii* GW-Imi-1b1. Samples were harvested at OD_550nm_ of 0.5 (t1: exponential phase), OD_550nm_ of 1 (t2: transient phase), OD_550nm_ of 2 (t3: early stationary phase), and OD_550nm_ of 3-4 (late stationary phase) in two independent replicates.

**Figure 3:**
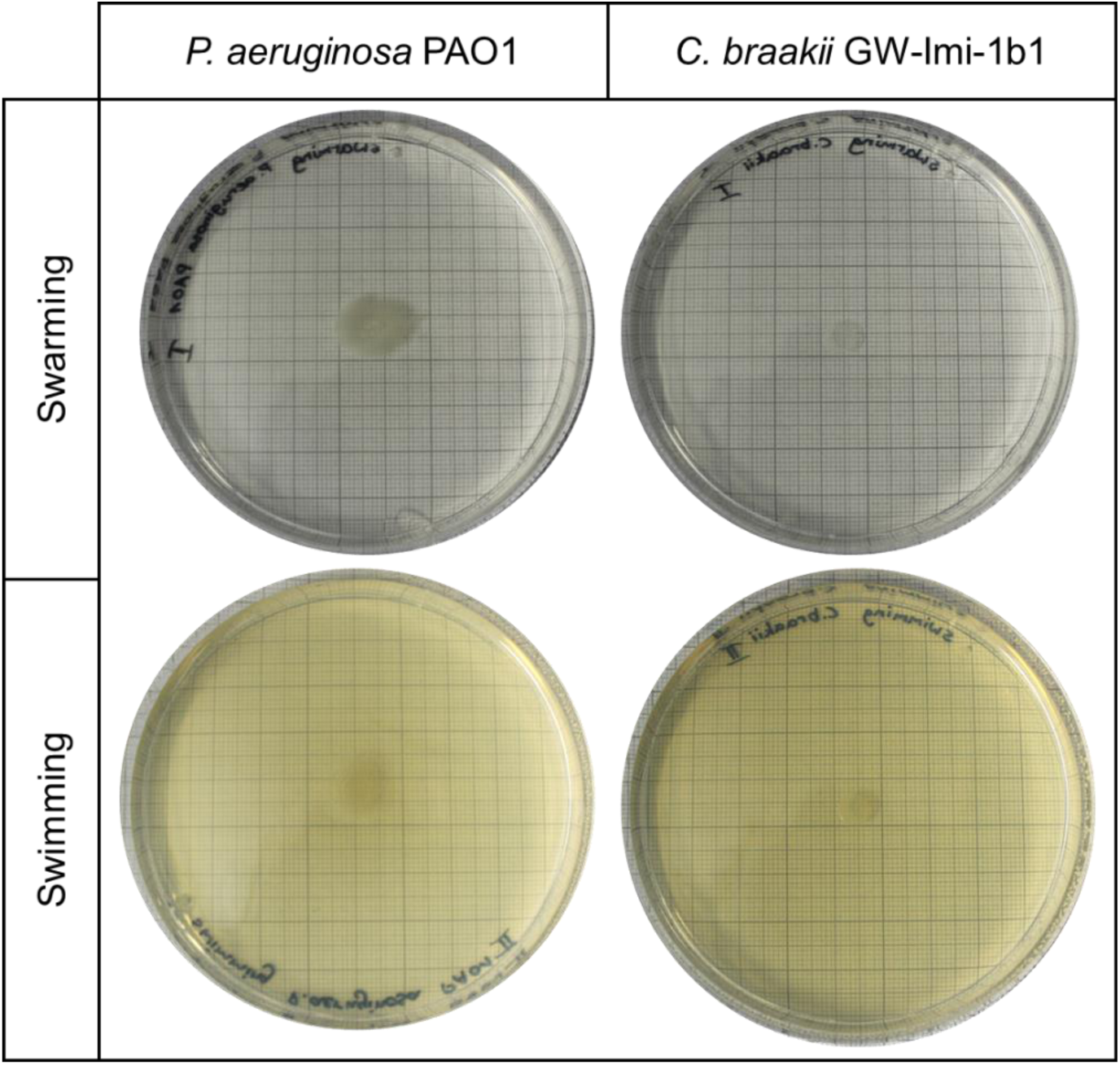
Test for swimming and swarming motility of *C. braakii* GW-Imi-1b1. *P. aeruginosa* PAO1 served as a positive control.

### Antibiotic resistance profile of *C. braakii*

The antibiotic resistance profile of *C. braakii* GW-Imi-1b1 was characterized using the disk diffusion method with a broad range of antibiotics. Ertapenem, imipenem, and meropenem were additionally tested using carbapenem E-tests. As summarized in Table 1, *C. braakii* GW-Imi-1b1 exhibits resistance to multiple antibiotics. While resistance to tobramycin was observed, the strain demonstrated susceptibility to the other aminoglycoside tested, amikacin. Cefotaxime demonstrates efficacy only at elevated concentrations, while the other cephalosporin, ceftadizime, exhibits no discernible activity. The fluoroquinolones ciprofloxacin, levofloxacin and ofloxacin demonstrated no effect on this strain. *C. braakii* GW-Imi-1b1demonstrates sensitivity only in the presence of elevated exposure to the carbapenems imipenem and meropenem, while exhibiting resistance to ertapenem. MIC was tested for these three carbapenems with an E-test. The average MIC for ertapenem is 12 µg/µl, which is considered as being resistant. As demonstrated by the disk diffusion method, the efficacy of the other two carbapenems is only evident at elevated concentrations. Specifically, meropenem demonstrates effectiveness at 2.5 µg/µl, while imipenem requires a concentration of 2.25 µg/µl for effective activity. According to the EUCAST guidelines, isolates are only susceptible at higher exposures for meropenem >2 µg/ml and imipenem >2 µg/ml (19). The supplemental material (**S1**) contains the images depicting the E-tests and the disk diffusion results.

**Table 1:** Antibiotic resistance profile of *C. braakii* GW-Imi-1b1. Standardized Kirby-Bauer test and EUCAST breakpoints were applied to determine susceptibility or resistance in two independent replicates.

| Antibiotic | *C. braakii* GW-Imi-1b1 | Inhibition zone [mm] |
| --- | --- | --- |
| Amikacin | susceptible | 28.5 |
| Amoxicillin + clavulanic acid | resistant | 12 |
| Aztreonam | susceptible, increased exposure | 25 |
| Cefotaxime | Susceptible, increased exposure | 18.5 |
| Ceftazidime | resistant | 14.5 |
| Chloramphenicol | resistant | 14 |
| Ciprofloxacin | resistant | 21.5 |
| Ertapenem | resistant | 12 |
| Imipenem | susceptible, increased exposure | 20.5 |
| Levofloxacin | resistant | 18 |
| Meropenem | susceptible, increased exposure | 17.5 |
| Ofloxacin | resistant | 13.5 |
| Tetracycline | resistant | 17.5 |
| Tobramycin | resistant | 12 |
| Trimethoprim/Sulfamethoxazol | susceptible | 29 |

### Proteome analyses of *C. braakii* GW-Imi-1b1

The proteome of the whole cell, as well as the extracellular proteome, were analyzed by LC-MS/MS. Approximately 2,360 proteins were found in the whole cell fraction and 1,300 in the extracellular fraction in at least two out of four replicates. Identification and quantification were conducted using the MaxQuant software, using a *C. braakii*-GW-Imi-1b1-specific database. The potential function of the identified proteins was determined by eggNOG-mapper version 2.1.9 (33) (Figure 4). Using this method, 217 proteins from the whole cell and 114 extracellular proteins including the hypothetical proteins could not be allocated. Nonetheless, this overview is useful for categorization and identification of proteins that might play a significant role in virulence. In Table 2, identified proteins associated with motility, secretion systems and antibiotic resistance are listed. With regard to the resistance-associated proteins, the following were identified: several β-lactamases, components of efflux pumps and transporters, and antibiotic-modifying enzymes were expressed by *C. braakii*-GW-Imi-1b1. Additionally, a detailed analysis of the data reveals that the β-lactamase (Protein ID: 05428) is among the ten most abundant proteins in the whole cell proteome, despite the absence of antibiotic stress during growth of the bacterium. A comprehensive analysis of the genome of *C. braakii* GW-Imi-1b1 was conducted, which revealed the presence of the β-lactamase gene on plasmid CP115731. It was further identified as a carbapenem-hydrolyzing class A β-lactamase (9). For this reason, we focused on this protein in further proteome analyses.

**Figure 4:**
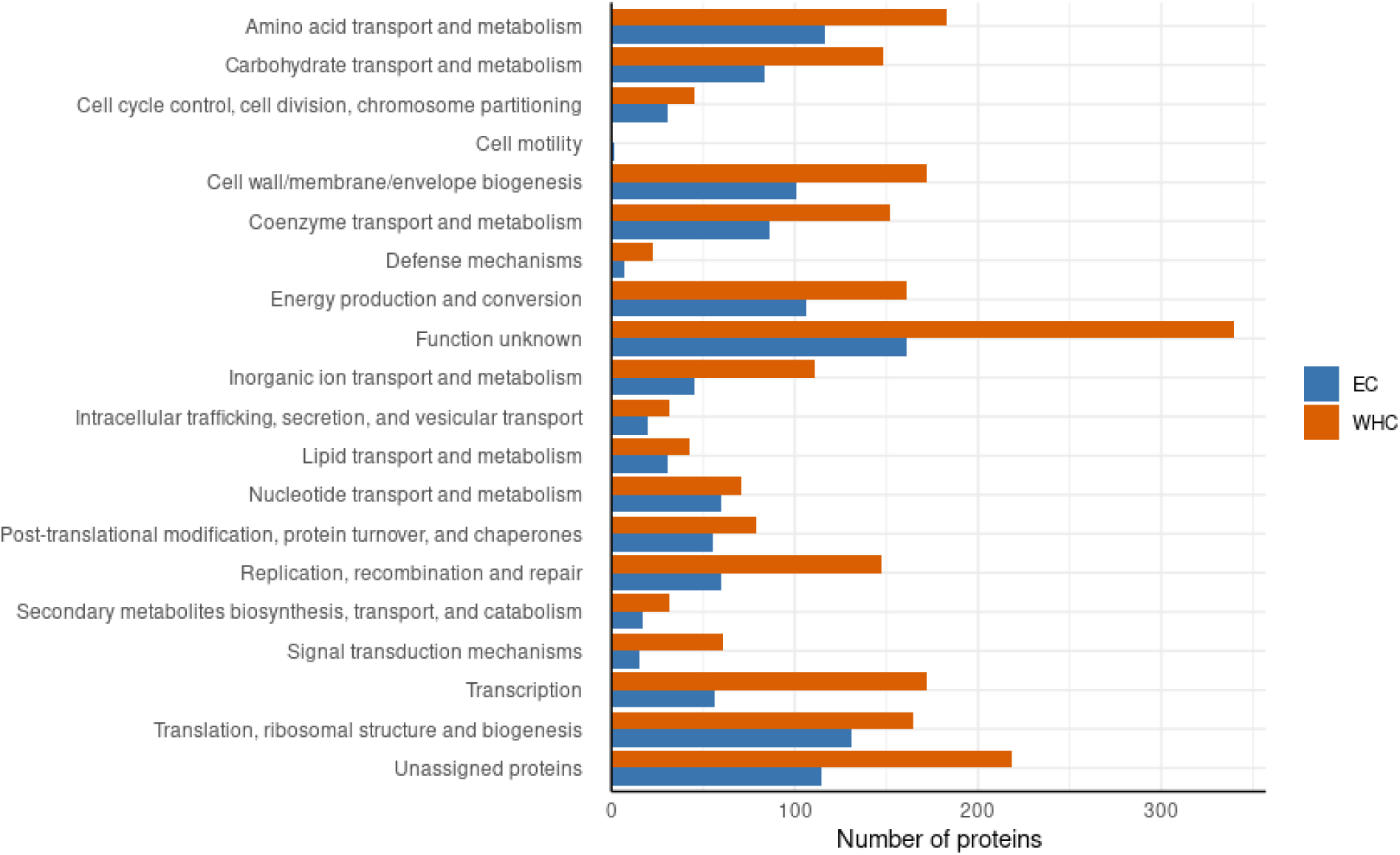
Functional annotation of the identified proteins by eggNOG-mapper. Whole cellular proteins (WHC) are marked in orange, extracellular proteins (EC) in blue.

**Table 2:**
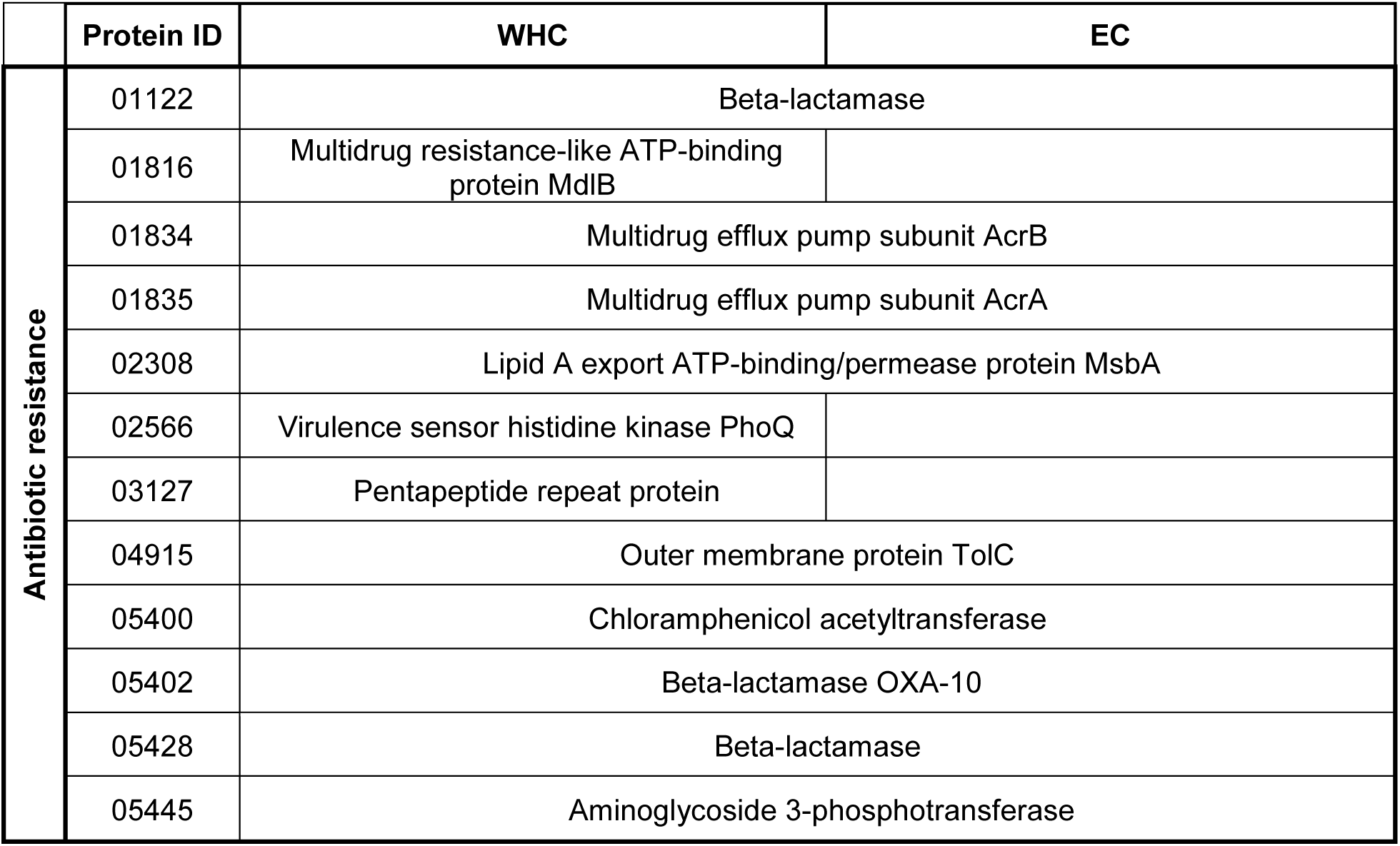
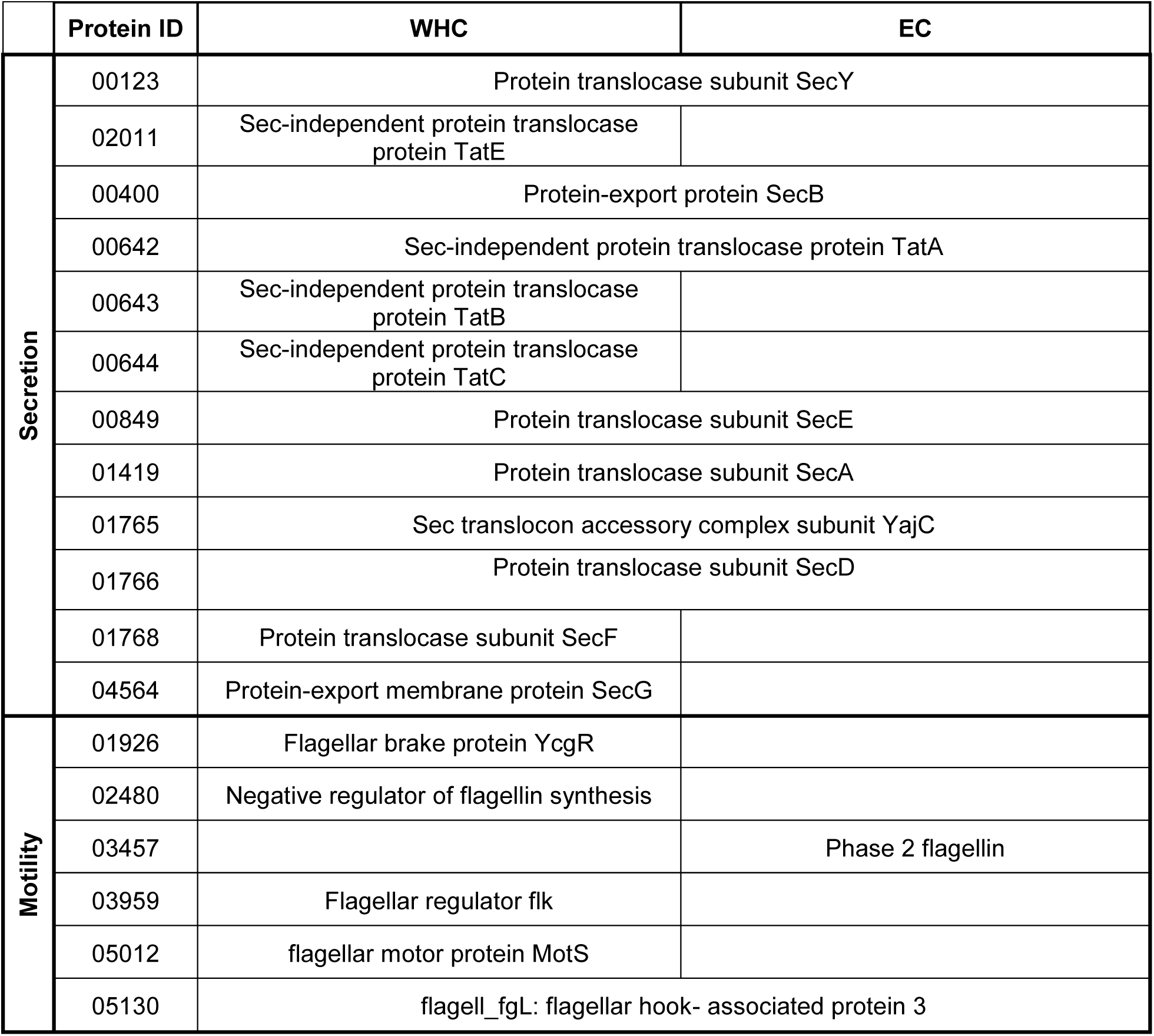
Selected proteins identified in the whole cell proteome (WHC) and extracellular proteome (EC) with respect to antibiotic resistance, motility and secretion.

### Effect of meropenem on *C. braakii* GW-Imi-1b1

As described above, the proteome profile exhibited an unusually high abundance of a β-lactamase. To test its activity, growth was determined about 26 h with different meropenem concentrations, ranging from 0.064 µg/ml to 20 µg/ml meropenem. As a comparison, *C. portucalensis* St35, a strain sensitive to meropenem, was tested as well. As shown in Figure 5, the growth curves between these two species differ significantly. While the growth of *C. braakii* GW-Imi-1b1 was almost not affected at all, already low concentrations of meropenem led to growth inhibition of *C. portucalensis* St35. Along this growth curve, samples for mass spectrometric analysis were harvested at the beginning from the pre culture(t0), after 2 h of incubation from start OD_500nm_ 0.1 (t1) and after 24 h (t2).

**Figure 5:**
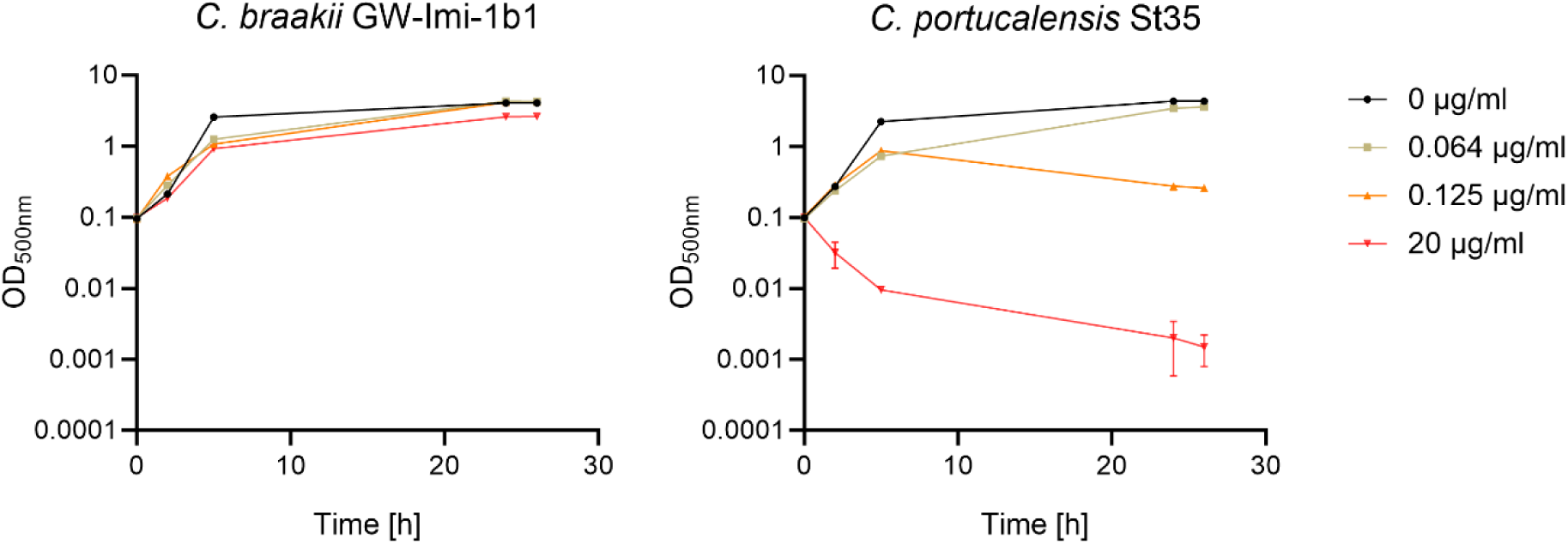
Growth curve of *C. braakii* GW-Imi-1b1 and *C. portucalensis* St35 in the presence of different meropenem concentrations.

The proteome was analyzed with a focus on the abundance of the aforementioned β-lactamase (ID: 05428). Independent of the presence or concentration of meropenem the specific β-lactamase was among the most abundant proteins in the cytosol (Figure 6).

**Figure 6:**
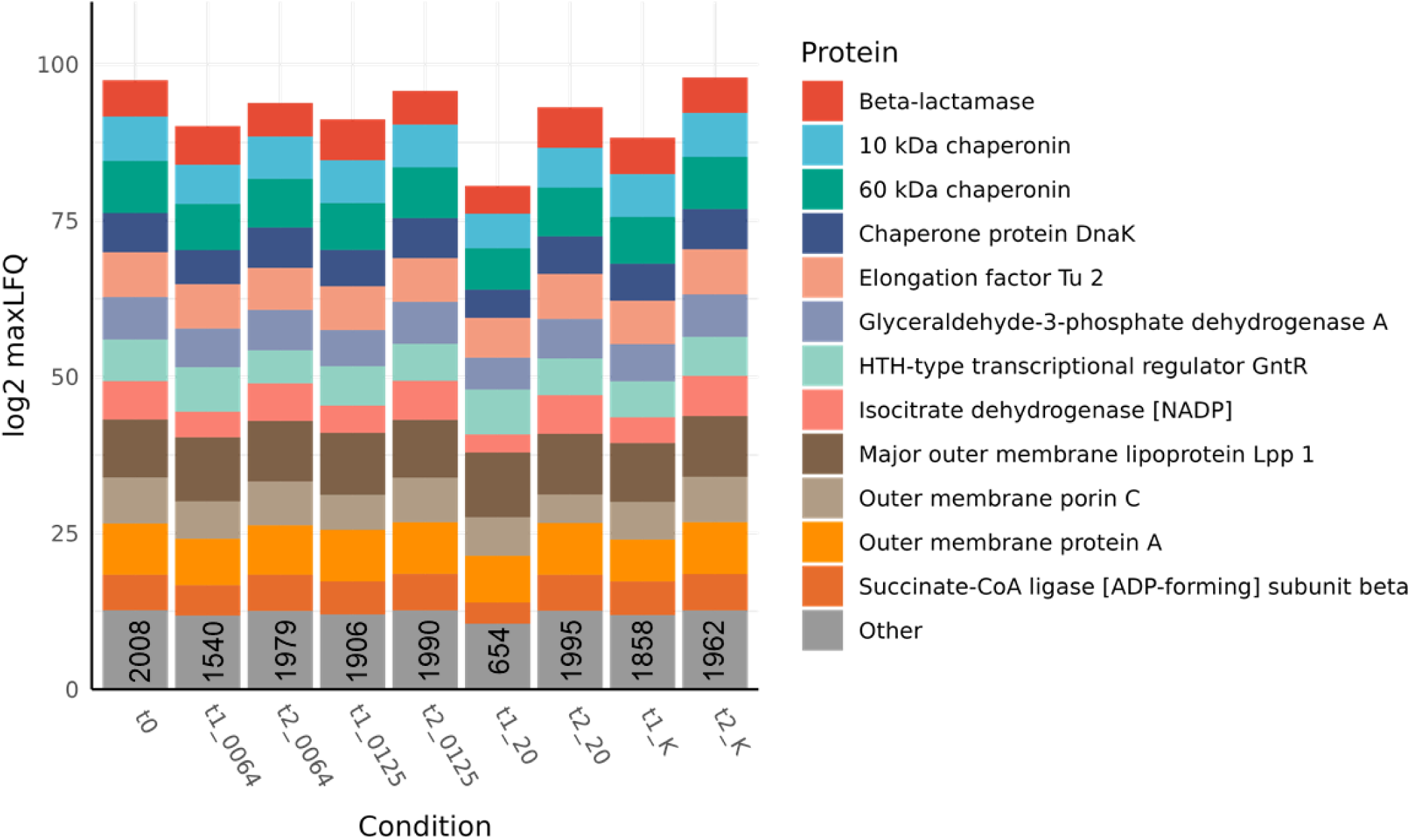
Abundance of the top 12 proteins in *C. braakii* GW-Imi-1b1. Normalized median of the maxLFQ of all proteins in each condition was calculated. The other proteins including the number are presented in grey. Samples were collected immediately before administration of the respective concentration of meropenem (t0), after 2 h (t1) and after 24 h (t2).

### The lytic phage vB_CbrP_HGW_001

#### Isolaton of a lytic phage for *C. braakii*-GW-Imi-1b1

The bacteriophage was isolated from raw sewage incubated with *C. braakii*-GW-Imi-1b1 as the host strain. On double agar overlay plates, after 18 h of incubation at 37°C, plaques with a size of 3-4 mm formed, that were surrounded by a halo.

#### Taxonomy and morphology of vB_CbrP_HGW_001

After isolation, vB_Cbr_HGW_001 was sequenced by Eurofins Genomics. The complete sequence of the vB_CbrP_HGW_001 genome comprises 39,389 base pairs (bp). Subsequent analysis with PHASTEST determined a total GC-content of 50.26 % and 44 open reading frames (Figure 7B). Transmission electron microscopy revealed the morphology of the isolated phage. As demonstrated in Figure 7A, vB_CbrP_HGW_001 is characterized by a short, non-contractile tail and an icosahedral capsid. The phage particles have a head diameter of 54 ± 3 nm and short tails of 11,8 ± 2,3 nm. Micrograph with a higher resolution is attached in figure **S2**. Consequently, it is regarded as being taxonomically classified within the *Caudoviricetes*. Furthermore, the significant alignments of vB_CbrP_HGW_001 with known phage genomes determined by BLASTN analysis suggest that it belongs to the genus *Kayfunavirus* (Table S1).

**Figure 7:**
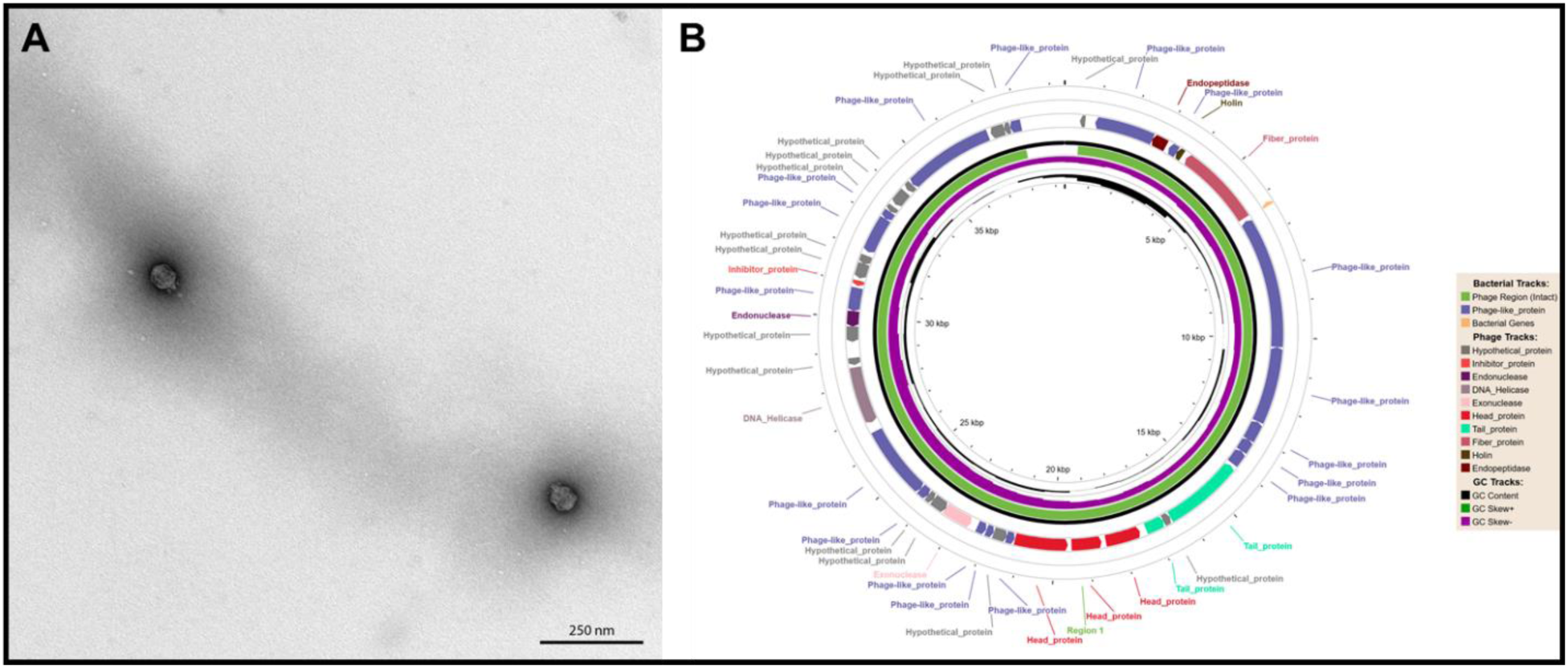
Morphological and genomic characterization of vB_CbrP_HGW_001. **A:** Transmission electron micrograph showing the phage particle with an icosahedral head and a short, non-contractile tail. **B:** DNA genome map with putative open reading frames (ORFs), color-coded by predicted functional categories as determined by PHASTEST analysis.

#### Properties of vB_CbrP_HGW_001

In order to provide further clarification regarding the properties of the isolated phage, a series of tests were conducted. In brief, the duration of an infection cycle was determined using adsorption kinetics. As shown in Figure 8A, titer of non-adsorbed phages after infection was determined. Almost immediately after infection, the phages were adsorbed, as almost no phage particles were detectable after 5 min. The replication of the phage, along with the subsequent lysis of the host cell, occurs within a period of 20 min, as indicated by the increase in the titer of non-adsorbed phages after this time duration. The burst size was subsequently computed to be 4 as depicted in Figure 8B. Stability of phage suspension was tested by incubation at various temperatures and pH values. A temperature of 4 °C and a pH value of 7.2 served as the controls, since the phage suspension (2.5×10^9^ PFU/ml) were stored under these conditions. Figure 8C revealed a decrease of PFUs at 50 °C and 60 °C. These findings indicate the phage retains stability over a temperature range of 4 °C to 40 °C. The test further indicated that a pH value ranging from 4 to 8 does not appear to exert an effect on the stability (Figure 8D). Even though a higher titer was calculated for pH 8 compared to the control, an unpaired t-test indicated no significant difference. However, even more acidic or basic pH values led to a clear decrease in the number of infectious phage particles.

**Figure 8.**
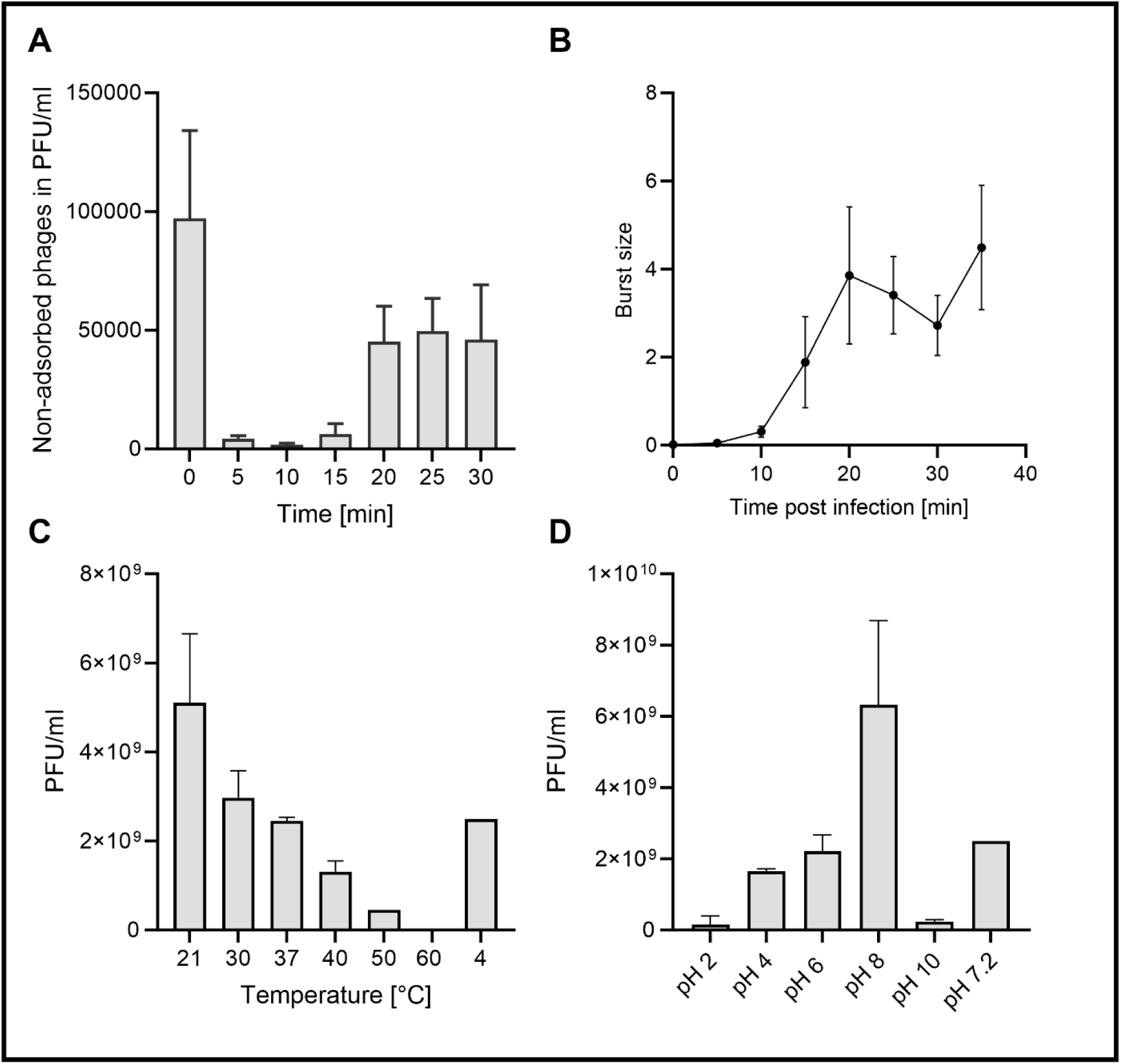
Properties of vB_CbrP_HGW_001. **A**: Adsorption kinetics of vB_CbrP_HGW_001. *C. braakii* GW-Imi-1b1 was infected with vB_CbrP_HGW_001 with an MOI=0.01 in the mid-exponential phase. **B:** Burst size of vB_CbrP_HGW_001. **C:** Temperature stability of vB_CbrP_HGW_001. **D**: pH stability of vB_CbrP_HGW_001. Phage suspensions were incubated at the respective pH value or temperature for 6 h. Titer was subsequent determined to observe phage stability.

## 4 Discussion

Multiresistant bacteria are a global threat to human health. In the context of healthcare facilities, there is an increased presence of pathogens that have developed resistances due to the widespread utilization of antibiotics. Moreover, these bacteria enter wastewater systems where they are capable of a horizontal transfer of resistance genes to other species (8,34). In this study, a carbapenem-resistant *Citrobacter* strain, isolated from hospital wastewater, was characterized in more detail. In addition, a lytic phage was found in the same environment, which could serve as a promising antimicrobial strategy alternative to antibiotic treatment.

The isolated *C. braakii* GW-Imi-1b1 exhibited several antibiotic resistances. Furthermore, there is a high potential for the production of OMVs, as evidenced by electron microscopy. OMVs were also reported in other Gram-negative pathogens like *Helicobacter pylori, P. aeruginosa,* and *Citrobacter rodentium* (35,36). Furthermore, they are associated with a variety of inflammatory diseases. As OMVs comprise lipopolysaccharides, peptidoglycans, and additional virulence factors, including enzymes and toxins, they are capable of triggering an immune response. However, in addition to their pathogen-associated function, OMVs have been attributed to interbacterial interactions (37). Therefore, *C. braakii* GW-Imi-1b1 may use its OMVs for either interbacterial interaction or virulence. Isolation and subsequent identification of the cargo is necessary to unravel the role of OMVs.

Proteases contribute to the virulence of a pathogen, i.e. degrading host cell membrane proteins and damaging tissues (38). Although *C. braakii* GW-Imi-1b1 expresses Tat- and Sec-secretion system, it does not secrete proteases at the tested conditions. Moreover, the mass spectrometric analysis identified only cytoplasmatic and periplasmatic proteases, a finding that aligns with the results of the protease assay.

The results of the motility test demonstrated that, in comparison to the positive control, *C. braakii* GW-Imi-1b1 exhibited a complete absence of motility with respect to swimming and swarming. In the course of proteome analysis, the presence of few flagellar proteins was confirmed. These flagellar proteins include “Phase 2 flagellin,” “Flagellar regulator flk,” “Flagellar hook-associated protein 1,” and “Flagellar motor protein MotS”. These are primarily structural components of flagella and proteins associated with the rotation motor. Flagella are especially needed for the purpose of swimming motility (39). Given the absence of motility observed in the assay, the suitability of the assay for *Citrobacter* is questionable. Further microscopic analyses, as transmission electron microscopy of *C. braakii* GW-Imi-1b1 could reveal the presence or absence of flagella (23). Notably, phase 2 flagellin was identified, suggesting two possible different antigenic phases for flagellin. In *Salmonella enterica* serovar Typhimurium, the expression of two distinct types of flagellin subunit proteins enables the pathogen to escape from the immune system and contributes to the virulence of this strain (40).

Several antibiotic resistances of *C. braakii* GW-Imi-1b1 were observed, which align with the results of the proteome analysis. All subunits comprising the AcrAB-TolC efflux system have been identified. This efflux system, composed of the membrane proteins AcrA, AcrB and TolC, has been demonstrated to contribute to resistance to multiple antibiotics by actively exporting them (41). Furthermore, a β-lactamase (Protein-ID:05428) was found in strikingly high abundance comparable with that of some ribosomal proteins. Moreover, sequencing of *C. braakii* GW-Imi-1b1 located the gene encoding for this β-lactamase on a plasmid. Hence, it could potentially be transferred by conjugation or transformation within the wastewater microbiome, thus transferring resistance to other organisms. Given the demonstrated carbapenem resistance and the unimpeded growth observed under varying meropenem concentrations, it is plausible that this β-lactamase contributes to the observed resistance. A proteome analysis of the carbapenem-sensitive strain *C. portucalensis* St35 revealed similar proteins involved in antibiotic resistance, but lacking the aforementioned β-lactamase (data not shown). The sensitivity of *C. portucalensis* St35 to meropenem underlines this hypothesis.

Overall, *C. braakii* GW-Imi-1b1 revealed different virulence factors like secretion systems, efflux pumps, motility proteins and production of OMVs. The treatment of this bacterium is particularly challenging due to its defense mechanisms, such as the high synthesis of β-lactamase conferring resistance to last resort antibiotics like carbapenems. The found lytic bacteriophage vBR_CbrP_HGW_001 is able to effectively lyse *C. braakii* GW-Imi-1b1 and could serve as a promising treatment. This bacteriophage was taxonomically classified within the *Caudoviricetes* with subsequent bioinformatically analysis to the genus *Kayfunavirus*. Stability assays revealed a notable environmental stability with respect to temperature and pH. A study by Liu *et al.* (42) reported another lytic bacteriophage belonging to the *Myoviridae* against another *C. braakii* strain. Similar to the phage in this study, capability of infection was affected in basic and acidic pH value and at temperatures higher than 50 °C. Furthermore, they exhibited synergistic effects of the bacteriophage with different antibiotics *in vitro* (42). It is reported for the treatment of enterococcal infections, that a combination of phage therapy and systemic antibiotics application can lead to fewer hospitalizations and to a reduction of the pathogen incidence in the patient’s stool (43). Future studies could thus focus on combining bacteriophages with antibiotics to reduce the occurrence of multiresistant bacteria in hospitals and wastewater.

## 5 Conclusion

In this study, a *Citrobacter* strain was isolated from hospital sewage from Greifswald, Germany. This multiresistant strain was further characterized and revealed a constitutively expressed β-lactamase, which was found to be amongst the ten most abundant proteins in the proteome. Moreover, this protein is potentially involved in the resistance to carbapenems, which complicates the treatment of this strain. In the same sewage, a lytic bacteriophage for this *Citrobacter* strain was found and characterized in detail. It could serve as a promising alternative treatment option against *C. braakii* GW-Imi-1b1. Further research is required to investigate a putative increased efficacy of combining this phage with antibiotics in a possible patient treatment scenario.

## Data Availability

All data produced in the present study are available upon reasonable request to the authors.

## Acknowledgements

This work was supported by a grant from the Helmholtz Association to Susanne Sievers.

